# Evaluating the clinical effectiveness and patient experience of an AI-based digital tool for home-based blood pressure management

**DOI:** 10.1101/2024.08.25.24312553

**Authors:** Alan Jelić, Igor Sesto, Luka Rotkvić, Luka Pavlović, Nikola Erceg, Nina Sesto, Zeljko Kraljevic, Joshua Au Yeung, Amos Folarin, Richard Dobson, Petroula Laiou

## Abstract

Hypertension, a prevalent cardiovascular condition, requires effective management of multimodal health risk factors. This study examines the effectiveness of a digital health tool designed for hypertension management and explores user perspectives on its utility. We analyse a cohort of 5,136 participants who used the digital tool, which provides continuous blood pressure monitoring, real-time feedback, and personalized health recommendations. Our results show that users achieve significant reduction in their blood pressure values and this reduction is positively correlated with the duration for which users report their blood pressure values. Additionally, we obtain high retention rates even after one year of using the digital tool. User feedback was collected through an online survey revealing high satisfaction rates. Participants highlighted the tool’s ease of use, and felt less anxious. Overall, our study demonstrates the potential of digital health tools in enhancing hypertension management and highlights the importance of user-centred design in developing effective health interventions.

## Introduction

According to the American Heart Association high blood pressure, called hypertension, is defined as systolic blood pressure (SBP) >= 130mm Hg or diastolic blood pressure (DBP) >=80mm Hg that remains elevated across time [Whelton_2018]. Hypertension affects over 1.2 billion people worldwide [WHO_2024] and in the last decades, the number of people with hypertension has doubled [Zhou_2021]. Hypertension is a major risk factor for cardiovascular diseases such as heart attack and heart failure, stroke, and chronic kidney disease [Chobanian_2003]. Hence, effective management of hypertension is a key factor in reducing morbidity and mortality associated with cardiovascular complications.

Traditional approaches to hypertension management usually require regular visits to healthcare facilities for blood pressure monitoring and consultations. Such approaches can be burdensome for patients and place a strain on healthcare systems. Moreover, patients commonly have difficulty with medication adherence and struggle to make lifestyle changes, with half of the patients stopping their treatment within a year [Public_Health_England_2017], this is a complex and multifactorial challenge. These factors result in poor blood pressure control leading to increased health risks, complications, mortality and hospital admissions. To address these health issues, there is a growing interest in leveraging artificial intelligence and digital health technologies (e.g., mobile apps, smartwatches) to facilitate home-based hypertension management. Such technologies offer long-term blood pressure monitoring, send reminders for the medication intake, provide educational content to support lifestyle changes and provide personalized health advice.

Recent trials have demonstrated that digital applications can significantly lower baseline 24-h ambulatory, home and office systolic BP compared with the control group [Nomura_2022, Kishi_2024]. Since 2022, there has been an explosion of interest in large language models (LLMs) for their conversational ability and their potential as clinical AI chatbots. Current LLMs built on the transformer architecture have demonstrated comprehensive medical knowledge [Singhal_2023], human-level empathy [Ayers_2023], and comparable bedside manner [Mukherjee_2024] and medical safety to clinicians. Implementation of an LLM-enhanced digital solution may improve user interaction and efficacy.

While the clinical benefits of long-term blood pressure monitoring with digital technology are well-documented [Bashshur_2016], it is of paramount importance to also understand the effectiveness for blood pressure reduction as well as perspectives of users for such digital tools. Investigating users’ perspectives for digital home-based blood pressure monitoring can contribute to the optimization and improvement of digital tools for hypertension as well as to ensure that such digital tools fulfil the needs and preferences of the users [Orji_2018].

In this work we analyse the effectiveness for blood pressure reduction, users retention and satisfaction with an innovative digital health tool that operates as a chatbot called Megi. Megi actively communicates with patients daily, reminds them to monitor their blood pressure, submit other relevant biometric data, take their medications, and enhance their lifestyle. Specifically, we analyse the retention and engagement of 5136 Megi users and investigate whether Megi users can achieve blood pressure reduction. Additionally, we present the results of an online survey that quantifies users’ perspectives about Megi. The insights obtained from this study can aid in the design, development and effectiveness of digital tools for hypertension and enhance users satisfaction.

## Methods

### Study design

We conducted an observational cohort study with 5136 Megi users who have been using Megi’s platform between June 2022 up to the end of July 2024 and fully completed the onboarding process. A user is considered fully onboarded once they accept the consent, enter their first name, last name, date of birth, and set the blood pressure measurement reminders. First, we analysed user engagement and retention on the platform across the whole user base to assess the platform’s overall success in retaining patients engaged and adherent with at-home blood pressure monitoring. We define retention rate as the percentage of users who continue to use Megi beyond specific time intervals after their initial use of the platform. In this study, we considered the intervals of 3, 6, 9, 12, and 24 months. Accordingly, users were grouped into cohorts based on the duration of their usage of Megi. These durations correspond to the specified retention periods. We analysed the retention based on two types of events: (1) blood pressure reporting, and (2) any interaction with Megi.

Subsequently, we randomly selected a cohort of active users that have been using Megi for at least three months to evaluate their perspectives about Megi. The randomly selected users had to fill an extensive online survey that consisted of 20 questions which were designed to (a) evaluate the usefulness of Megi in improving self-management of hypertension; (b) assess their satisfaction; (c) report their willingness to use Megi and (d) assess their behavioural response for Megi. The survey was conducted in June 2023 and all participants gave written informed consent that they agree to participate in the survey. Data was stored on a secure cloud platform that adheres to protected health information management standards. The study was approved by the Magdalena University Hospital Research Ethics Committee (195/Inf-701/22).

### Digital tool for the management of hypertension

All study participants used Megi, a digital tool for hypertension management. Megi is a chatbot equipped with a specialized transformer-based AI model, and is accessed via messaging apps such as WhatsApp and Viber. Via this chat and through images, Megi gathers on a daily basis multimodal behavioural and biometric data of the users that range from blood pressure, and heart rate measurements to mood and stress levels. Additionally, it provides personalized advice for lifestyle changes. On a daily basis, users receive reminders to measure their blood pressure and take their medications. All data are stored in secured cloud platforms and clinicians have 24/7 access to all data via an online platform.

### Survey questions

The online survey consisted of 19 questions. There were three questions that collected information about the users age, sex and employment status. In addition, there was one question that evaluated the impact of hypertension on the users daily life. The survey had three questions that assessed the usefulness of the technology in improving self-management of hypertension (i.e., “I understand why it is important to measure your blood pressure daily”; “I would often forget to measure my blood pressure if Megi didn’t remind me”; “With Megi, I will develop the habit of regular blood pressure measurement and monitoring”). Moreover, there were three questions that evaluated the users satisfaction (i.e., “I can see the benefits of using Megi”; “I have confidence in Megi”; “I would continue to use Megi even if my doctor did not access my data”). The willingness to use the digital tool was evaluated by 4 questions (i.e.; “I would definitely recommend Megi to my friends”; “Anyone who cares about their health should use Megi”; “For the sake of myself and my loved ones, it is important for me to take care of my health”; “My loved ones and friends would encourage me to use Megi”), whilst the behavioural response of the users was also assessed by 4 questions (i.e., “Since using Megi, I feel safer and more relaxed”; “I feel happy when I receive messages from Megi”; “Megi makes me feel less lonely”; “I wouldn’t want to lose access to Megi”). Finally the survey had one question that asked users to provide the unmet needs of the digital tool in free-text. The answers were provided either in a 5-point Likert scale (1: I strongly disagree, 2: disagree, 3: neither agree nor disagree, 4: agree, 5: strongly agree) or in free text.

### Statistical analysis

We analyzed the blood pressure values of all users in the first two weeks after their enrolment to the MEGI platform and in the last two weeks before they stopped reporting their blood pressure values. For every user we computed the average blood pressure values in the first and last biweekly segment. To quantify the statistical differences between the first and last biweekly segment we applied a one-sided non-parametric Wilcoxon rank-sum test. Results were considered statistically significant if their p-value was less than 0.05. Additionally, we performed a descriptive analysis to assess the results of the online survey. Informative quotes were provided for the answers that were given in free text. All analysis was performed in MATLAB R2022a.

## Results

### Users Retention

We first analysed the retention of 5136 Megi users (2399 female; 2733 male; 4 other; median age: 59 years; age range 17-95 years). Retention was quantified based on (a) blood pressure reporting, and (b) any interaction with Megi. Figure 1 demonstrates that the retention rates after one year of using Megi is 50.3% for users based on all their interactions with Megi (e.g., updating weight data, reacting to therapy reminders, submitting feedback) and 48.1% for users based on their blood pressure measurement reports. The retention rates are even higher i.e., 60.5% and 63.1% respectively for users who self-reported to be hypertensive. Interestingly, user retention remains stable even after 2 years of using Megi.

**Figure 1:**
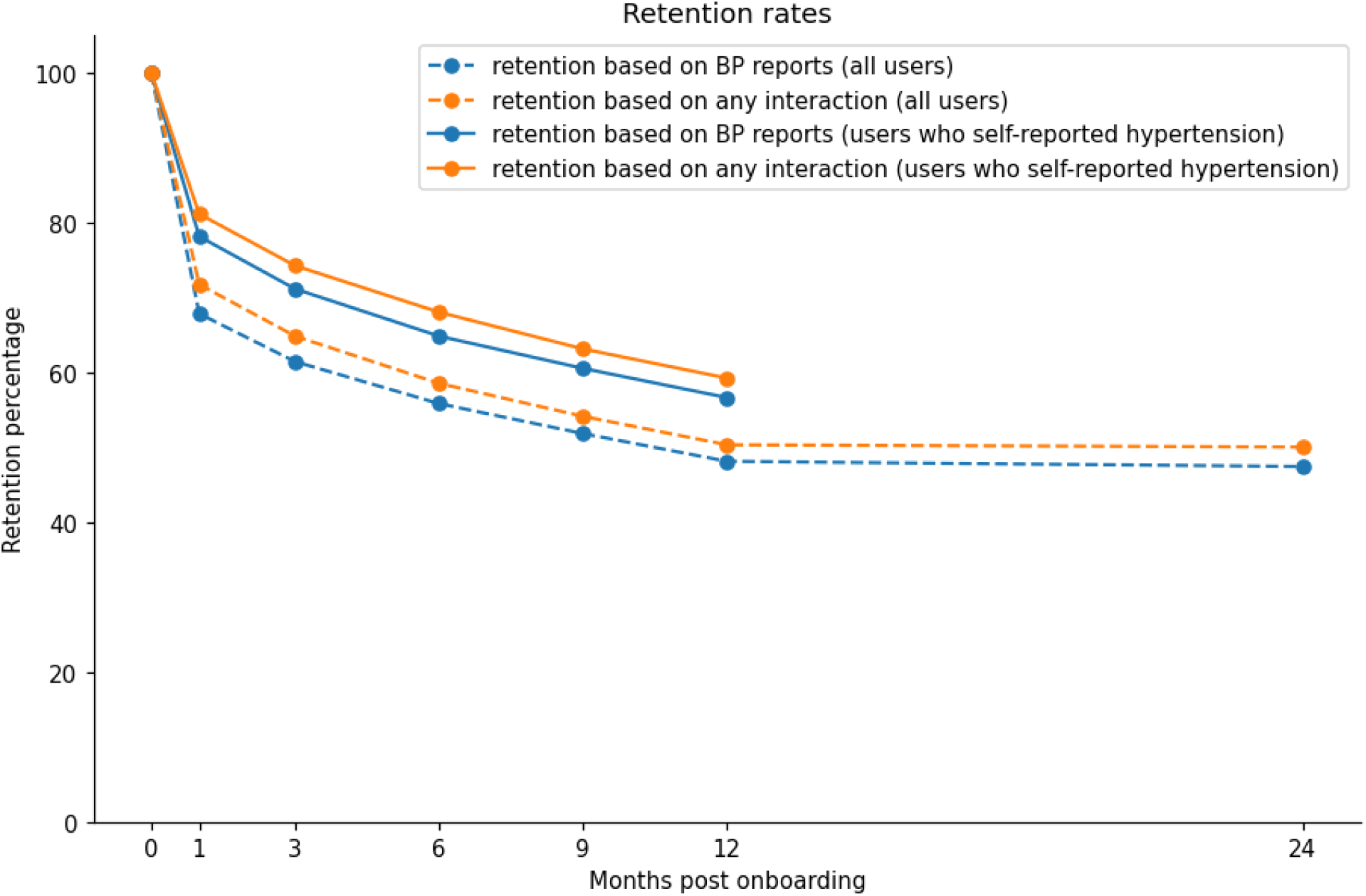
Users retention (n=5136) across months post-onboarding based on blood-pressure reporting (orange) and any interaction with Megi (blue). Dotted lines correspond to all users, whilst solid lines correspond to users who self-reported that have hypertension upon their enrolment (this feature was implemented in the second year).

### Blood pressure analysis

4736 of 5136 (92.2%) users reported their blood pressure values. We first seek to uncover whether there is a statically significant difference in the average systolic blood pressure (SBP) measurements between the first and last biweekly segment. Figure 2 illustrates the distributions of the average SBP values of all users for the first 14 days after their enrolment and last biweekly segment for which users reported their blood pressure values. We applied an one-sided Wilcoxon rank sum test and found that the SBP values in the last biweekly segment were statistically significantly reduced compared to smaller than the first biweekly interval (p-value: 0.0002).

**Figure 2:**
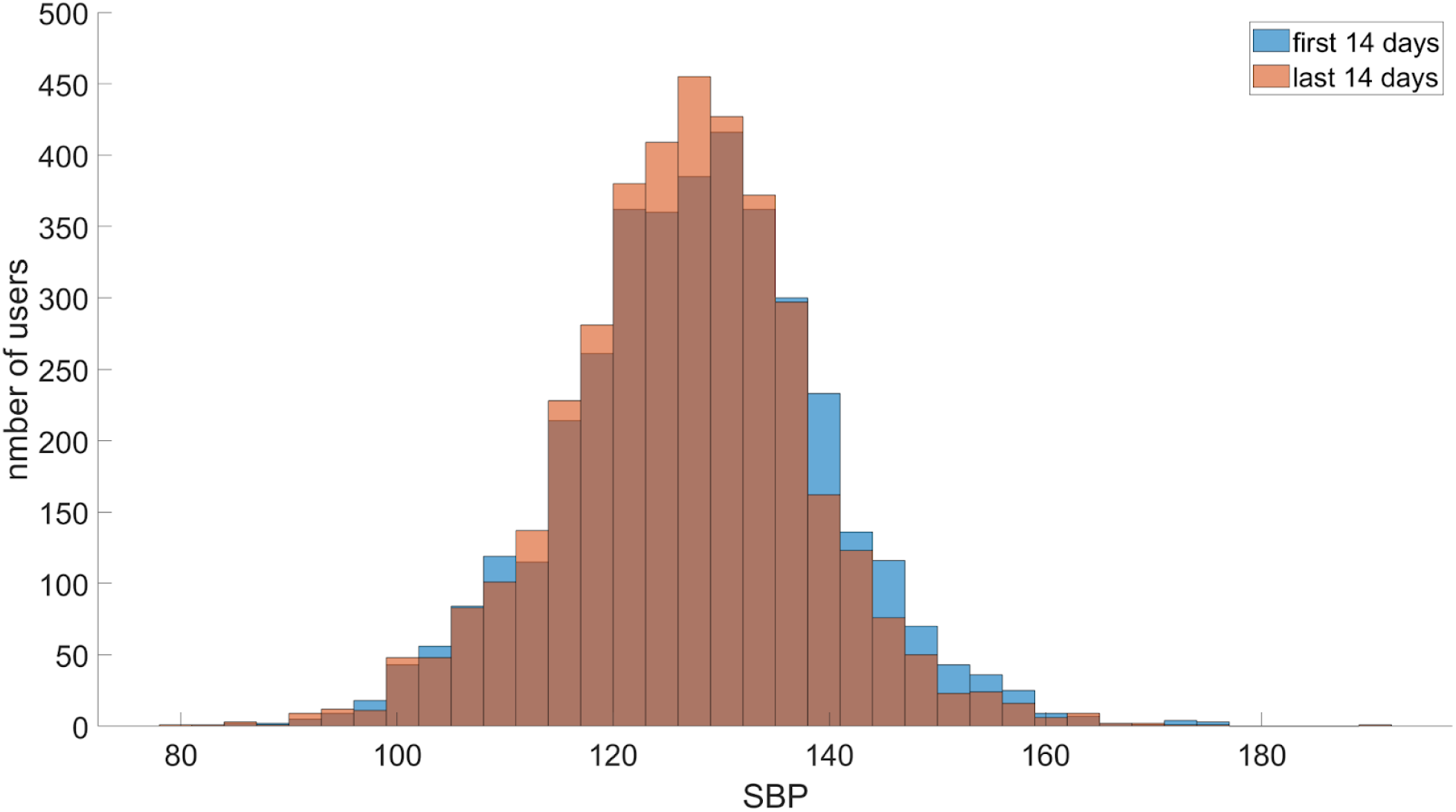
Distributions of the average systolic blood pressure measurements during the first biweekly segment after study enrollment (blue) and last biweekly interval (orange) before users paused reporting blood pressure values.

Afterwards, we wanted to explore whether such SBP differences were stronger for users whose average SBP values were at least 140mmHg in the first biweekly segment (Figure 3). We saw that such differences between the first and last biweekly interval were much stronger (p-value: 1.7 x10^-9)(Figure 3a). We also examined the correlation between the SBP drop (i.e. the difference of the average SBP during the first biweekly interval and the last biweekly interval) and the period for which users report their blood pressure values. The Spearman correlation was 0.53 and it was statistically significant (p-value = 4.5^-32) which means that larger SBP reduction is positively correlated with longer usage of Megi (Fig. 3b). Figure 3c illustrates the SBP drop across the months for which users were reported their blood pressure values. The maximum duration in blood pressure reporting was 24 months and we observe that there is an increasing trend of the SBP drop across the time (Fig. 3c).

**Figure 3.**
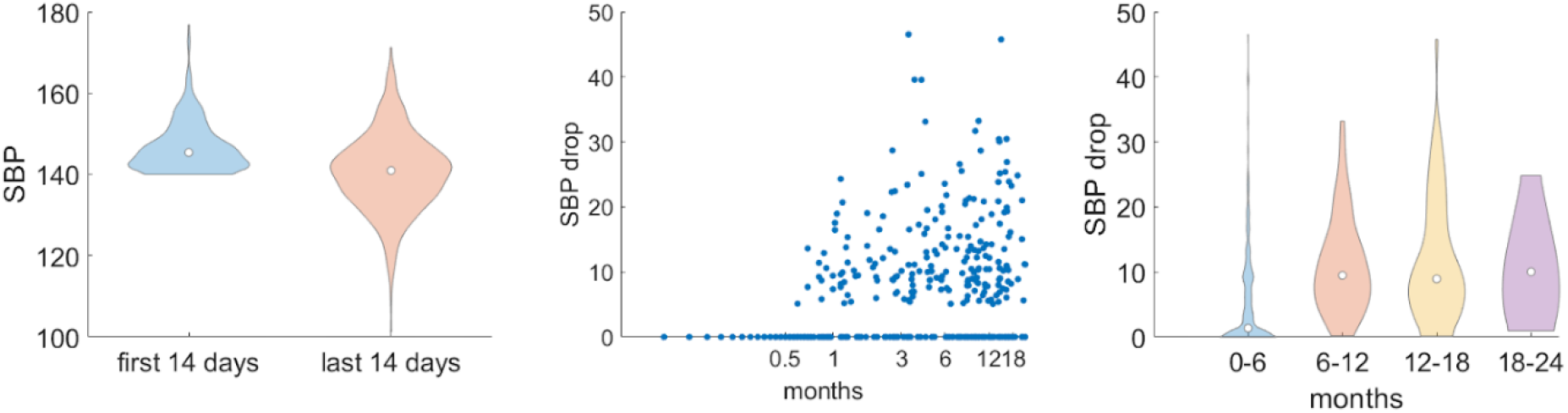
(a) Distributions of SBP measurements for users whose average SBP value was at least 140mmHg during the first weekly biweekly interval after enrolment (blue). The orange violin plot depicts the average SBP values during the last biweekly interval for which users reported their blood pressure values. (b) SBP drop across the time (in months) for which users report their BP measurements. Each dot corresponds to a different user. (c) Distributions of the SBP drop across different time intervals (in months) for which users report the blood pressure measurements.

We performed a similar analysis for the users whose average SBP was at least 150mmHg in the first biweekly segment (Figure 4). For users with such high blood pressure the differences between the first and last biweekly interval were also strong (p-value: 1.7 x10^-9) and the correlation between the SBP drop and the duration for which users report their blood pressure measurements was even stronger (cor:0.75; Figure 4b). This finding is also confirmed from Figure 4c that illustrates clear increase in the SBP drop across the time.

**Figure 4.**
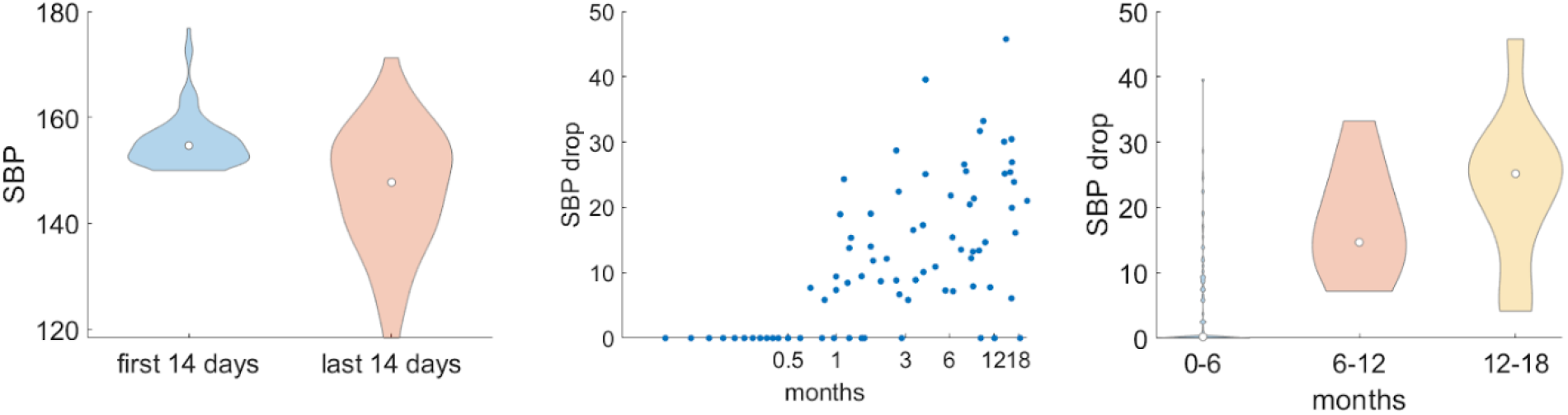
Similar to figure 4 but for users whose average systolic blood pressure was at least 150mmHg during the first biweekly interval after their enrolment.

### Online survey analysis Dataset characteristics

A total of 129 people clicked on a link to access the survey (June 2023). The total number of analysed participants is 125 (3 participants were excluded from the analysis due to the presence of missing values). Summary demographics regarding participants age, sex and employment status are provided in Table 1 and Figure 1. Out of the 125 participants 50 were females and the age of all survey participants ranged from 23 to 82 years (mean: 60; standard deviation: 11.6).

**Table 1:** Dataset characteristics.

| Characteristics | Number of users (N, %) |
| --- | --- |
| Age group |  |
| [20-29] | 2 (1.6) |
| [30-39] | 3 (2.4) |
| [40-49] | 18 (14.4) |
| [50-59] | 27 (21.6) |
| [60-69] | 52 (41.6) |
| [70-79] | 20 (16.0) |
| [80-89] | 1 (0.8) |
| Sex |  |
| Male | 70 (44.0) |
| Female | 55 (56.0) |
| Employment status |  |
| Employed | 54 (43.2) |
| Unemployed | 5 (4.0) |
| Retired | 64 (51.2) |
| Student | 1 (0.8) |

### Impact of hypertension in the responders daily life

At first study participants were asked whether high blood pressure has a significant impact on their life. The vast majority of the participants strongly agreed (47%) or agreed (29%) whilst just 19% of them strongly disagreed or disagreed (Fig. 6).

**Figure 5:**
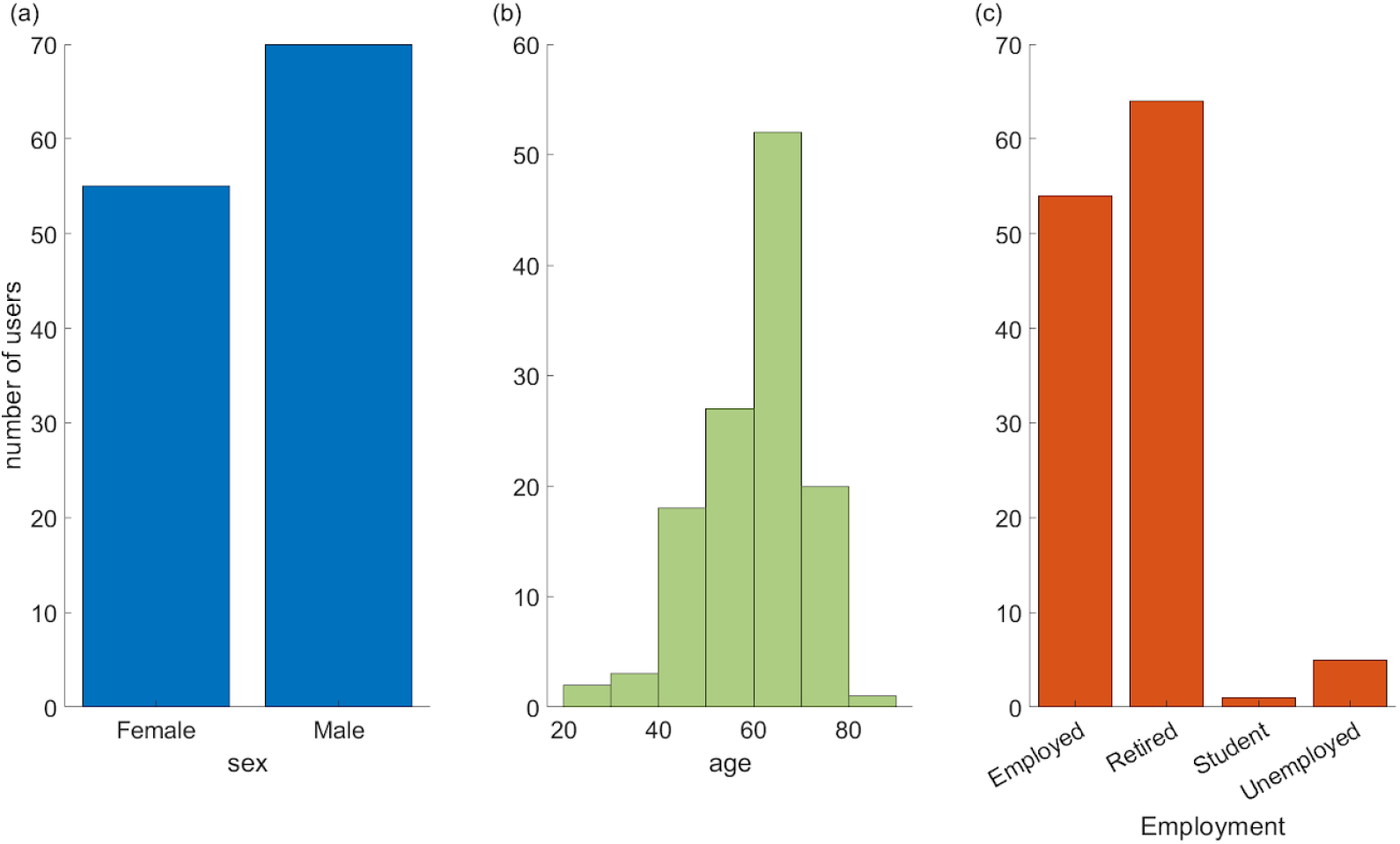
Descriptive characteristics of sex (a), age (b) and employment status (c).

**Figure 6:**
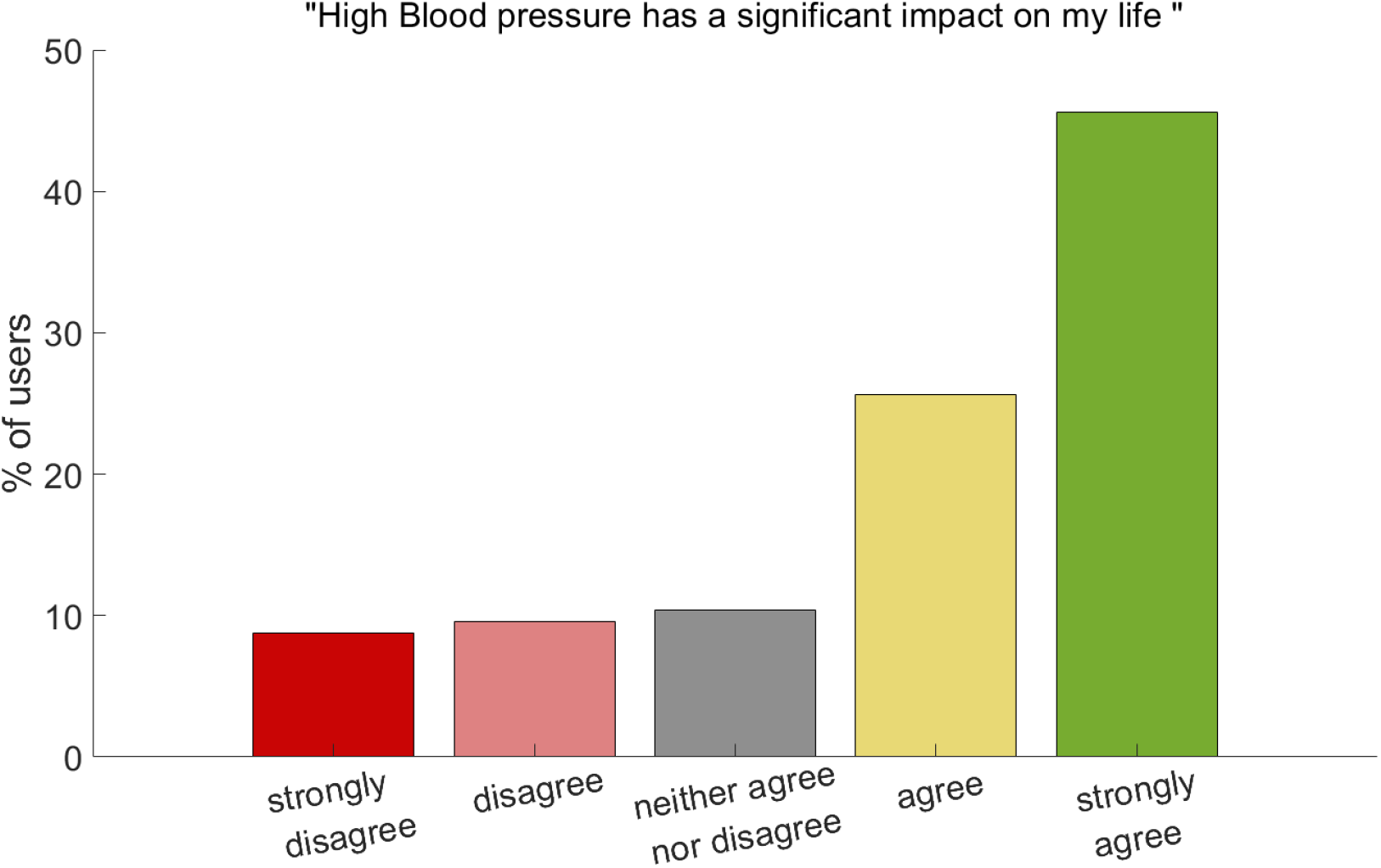
Impact of hypertension on users’ daily life

### Usefulness of technology in improving self-management

Participants were asked a series of questions to evaluate the usefulness of technology in managing their hypertension. The responses indicated a high level of perceived usefulness. Overall, more than 80% of the participants agreed or strongly agreed that Megi helped them understand their hypertension better. Regarding the importance of blood pressure monitoring 98% found the technology useful for regularly monitoring their blood pressure (Fig. 7a). When participants asked whether they would forget to measure their blood pressure if MEGI did not remind them, more than 80% responded positively (Fig 7b). In addition, study participants felt that using MEGI makes them develop the habit to measure their blood pressure regularly (more than 95% responded positively; Fig. 7c)

**Figure 7:**
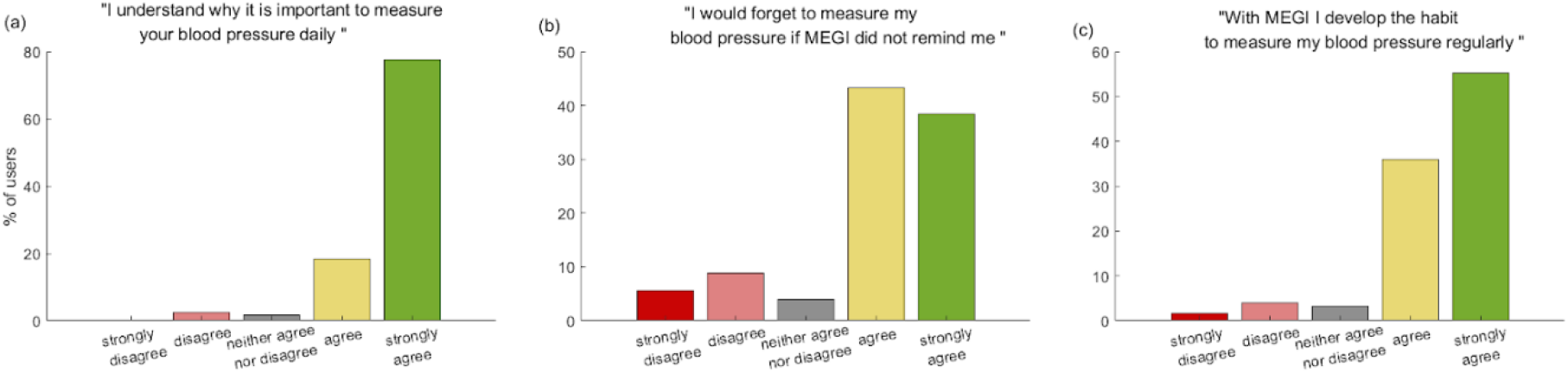
Usefulness of technology in improving self-management

### User satisfaction

When participants answered questions that assessed their overall perception, confidence in the digital tool, and willingness to continue using it independently of their healthcare provider again their overall satisfaction was very positive. Specifically, regarding the statement “I can see the benefits of using Megi.”, 98% of study participants strongly agreed or disagreed (Fig. 8a). In addition, when participants asked whether they have confidence in MEGI 92% responded positively (Fig. 8b), whilst when they asked whether they have the willingness to use MEGI independently more than 80% agreed or strongly agreed (Fig. 8c).

**Figure 8:**
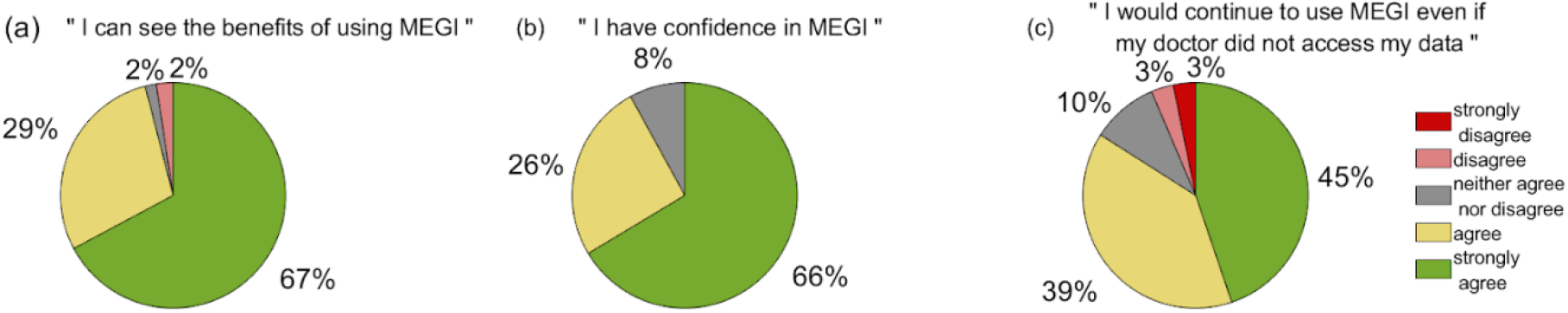
User satisfaction about technology

### Willingness to use the technology

Study participants were also assessed regarding their willingness to use or recommend MEGI. Specifically, they asked whether they a) would recommend Megi to their friends (more that 90% of the users responded positively) b) agree with the statements i) “anyone who cares about their health should use Megi” (more that 80% of the users strongly agreed or agreed); c) “For the sake of myself and my loved ones, it is important for me to take care of my health” (more that 90% of the users responded positively) and d) “My loved ones and friends would encourage me to use Megi” (more that 55% of the users responded positively). Again in all questions of this category

### Behavioral response

At the end of the survey participants had to answer questions regarding their behavioural response with the digital tool. They had to answer the following questions: Since using Megi, I feel safer and more relaxed; I feel happy when I receive messages from Megi; I wouldn’t want to lose access to Megi; Megi makes me feel less lonely. Across all questions (Fig. 10a-c) but the last (Fig 10d) more than 80% of the participants felt that MEGI reduces their anxiety and feel safe.

**Figure 9:**
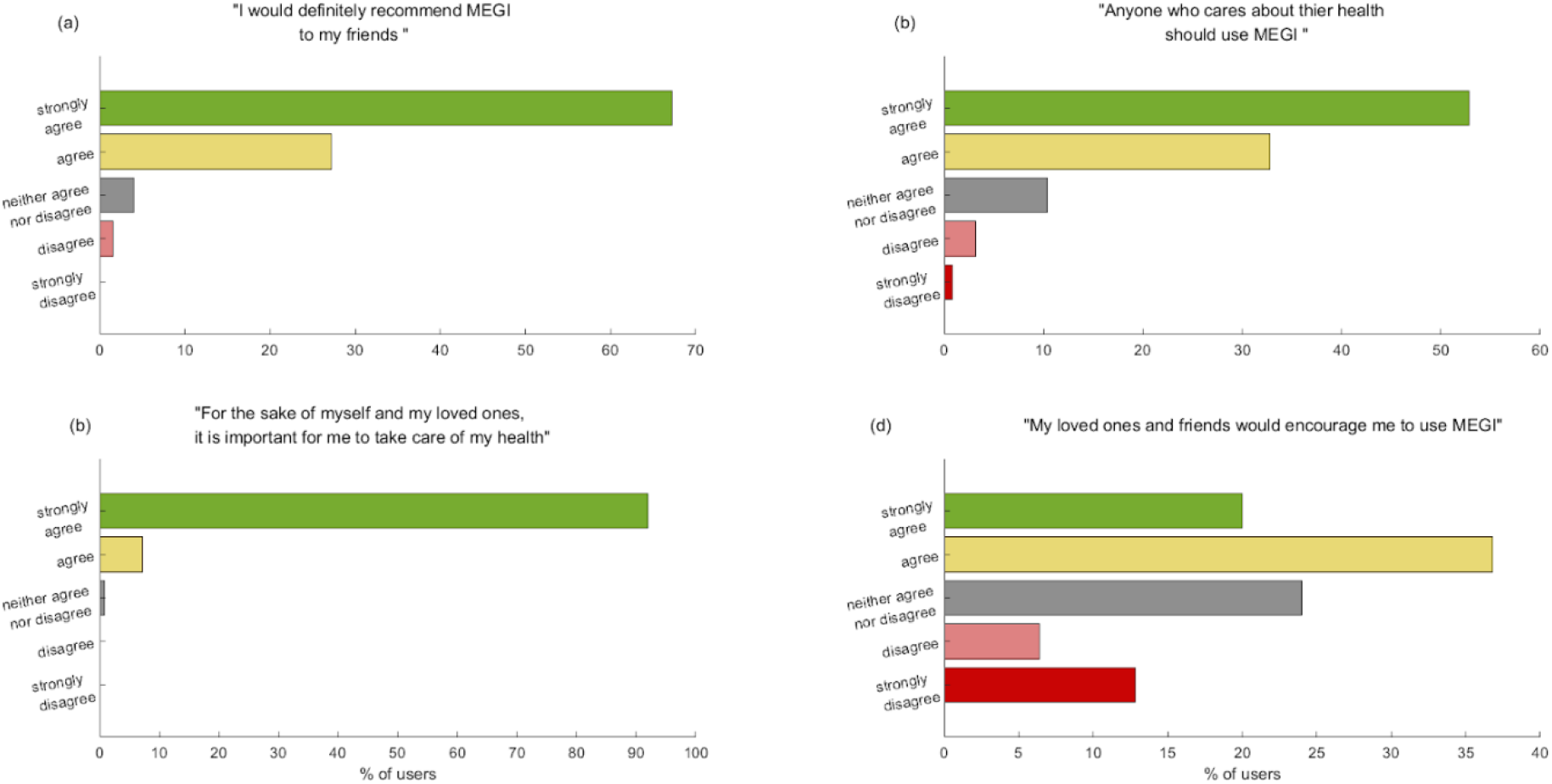
Users’ willingness to use or recommend technology.

**Figure 10:**
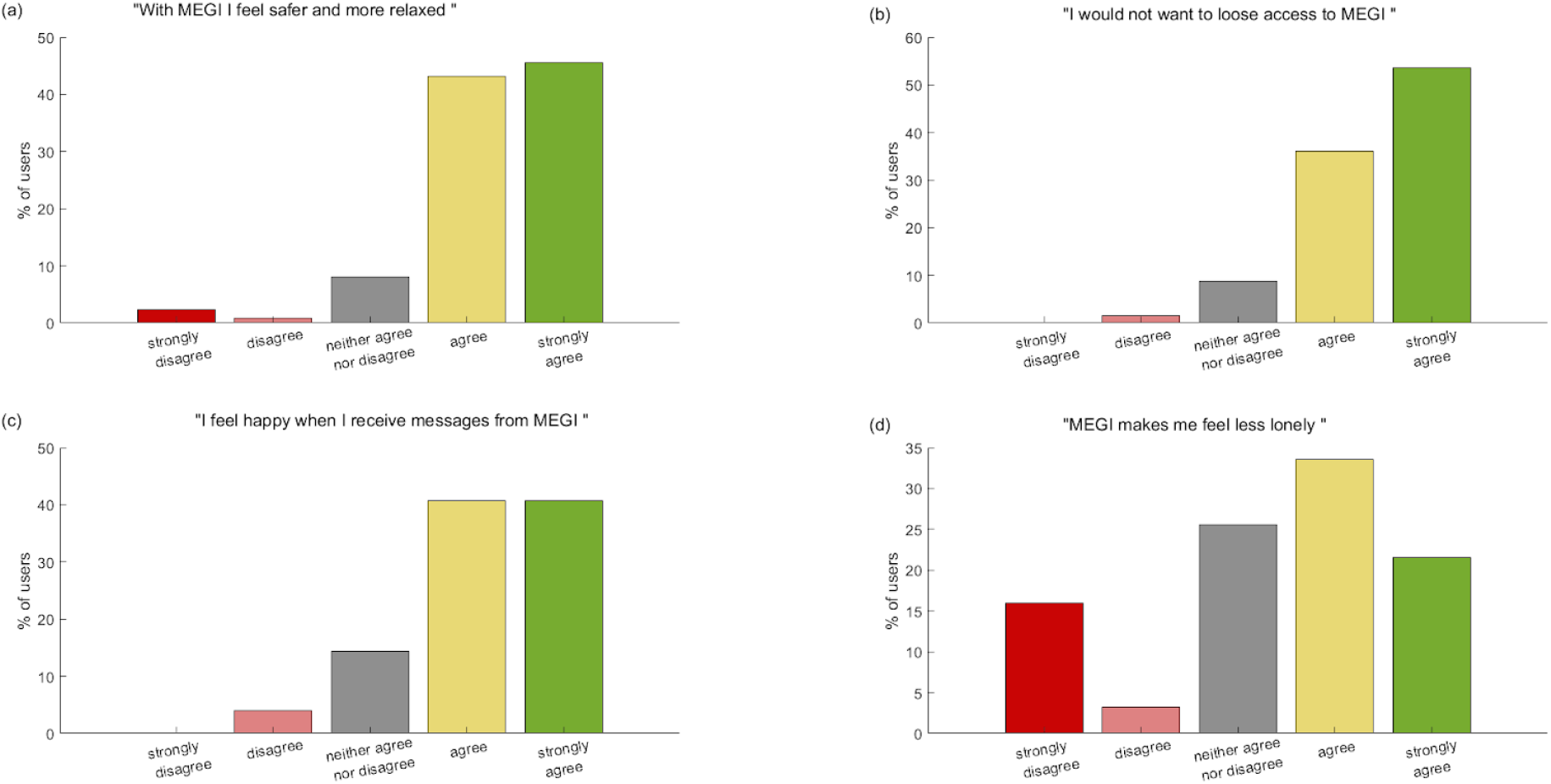
Users behavioral response about technology

### Unmet needs

Study participants were also asked whether there is something that they would change about Megi. Answers were given in free-text. Some users were asking for more personalized treatment (e.g.”I think maybe she should adjust according to the specifics of individual patients. I can illustrate this with my own example. I have been hypertensive for about thirty years, I regularly measure my blood pressure, so I don’t think it’s necessary to report my condition every two weeks.”; “I would like to determine more precisely what type of pressure I have… when it is really low and when it is high, and based on the measurements…”) or the possibility of clinical consultations with cardiologists (e.g. “Maybe the availability of a cardiologist to guide me”; “If there were an opportunity to occasionally discuss the results with a doctor, I think I would take regular measurements more seriously.”)

## Discussion

The management of hypertension at home using digital apps presents a promising avenue for improving patient outcomes and alleviating the burden on healthcare systems. The present study explored users’ perspectives on Megi, a digital tool designed to facilitate long-term home-based blood pressure monitoring and demonstrated the effectiveness of Megi on blood pressure reduction.

Analyzing the activity of 5136 users, our results show very high engagement and adherence to blood pressure readings that remain stable even after one year of using Megi (Figure 1). Specifically, more than 50% of users are active on the Megi digital health platform for 12 months, which demonstrates higher user retention when compared to previously reported industry standards. For example, consistent with other large-scale digital health studies, the notable Stanford-led MyHeart Counts study experienced substantial dropout rates; mean engagement with the app was only 4.1 days. (Hershman_2019). <u>Statista’s</u> research into worldwide retention rate of mobile app installs reveals that on the day of installation, the average retention rate across the 31 categories was 25.3%. Worryingly, by day 30, this percentage drops to just 5.7% and when it comes to the Health and Wellness industry specifically, even lower to 2.8% (Statista_2023).

In addition, our analysis showed the effectiveness of Megi as a digital tool for hypertension management (Figures 2-4). We demonstrated that the average SBP of the last biweekly interval for which users reported their blood pressure values was statistically significantly smaller compared to the first biweekly segment after users’ enrolment (Figure 2). This difference was even stronger for users who had high (>140mmHg; Figure 3a) and very high SBP measurements (>150mmHg; Figure 4a) during the first biweekly segment. Furthermore, we showed that there is a positive correlation between the SBP drop and the duration for which users report their blood pressure values. This means that larger SBP reductions are analogous with longer usage of Megi (Figs 3,4b,c)

Through an online survey, we evaluated users’ perspectives on the technology’s usefulness in improving self-management of hypertension (Figure 7), their satisfaction with the tool (Figure 8), their willingness to use it (Figure 9), and their behavioural responses to its implementation (Figure 10). The findings indicate a high level (i.e. more than 90%) of user satisfaction with Megi. Participants strongly acknowledged the technology’s usefulness in enhancing their ability to manage hypertension independently (Fig. 3,4), as well as they expressed their willingness to suggest Megi to other users (Fig. 5). The tool’s integration into daily routines led to a noticeable reduction in anxiety and an increase in feelings of relaxation and happiness (Fig.6), suggesting a positive impact on mental well-being. Furthermore, the willingness of users to recommend Megi to friends reflects strong approval and confidence in its efficacy. Overall, the positive feedback from users highlights the potential of Megi as a valuable digital tool for hypertension management.

Despite the valuable insights, this study has limitations that must be acknowledged. The single ethnicity of the analysed cohort limits the generalizability of the findings. Additionally, the evaluation focused exclusively on one digital tool, e., Megi, which may not provide a comprehensive understanding of the broader landscape of digital tools for hypertension management. These constraints may affect the generalizability of the findings and highlight the need for further research with more diverse and larger populations to validate and expand upon these results.

Our study demonstrated the effectiveness of a digital tool for hypertension management and highlighted the importance of understanding user views in the design and implementation of digital health interventions for hypertension management. In conclusion, the insights obtained from this study highlight the potential of digital apps to enhance hypertension management by providing continuous monitoring, real-time feedback, and personalized health recommendations.

## Data Availability

No data are available.

